# Improving Coronavirus (COVID-19) Diagnosis using Deep Transfer Learning

**DOI:** 10.1101/2020.04.11.20054643

**Authors:** Arshia Rehman, Saeeda Naz, Ahmed Khan, Ahmad Zaib, Imran Razzak

## Abstract

**Background:** Coronavirus disease (COVID-19) is an infectious disease caused by a new virus. Exponential growth is not only threatening lives, but also impacting businesses and disrupting travel around the world.

**Aim:** The aim of this work is to develop an efficient diagnosis of COVID-19 disease by differentiating it from viral pneumonia, bacterial pneumonia and healthy cases using deep learning techniques.

**Method:** In this work, we have used pre-trained knowledge to improve the diagnostic performance using transfer learning techniques and compared the performance different CNN architectures.

**Results:** Evaluation results using K-fold (10) showed that we have achieved state of the art performance with overall accuracy of **98.75%** on the perspective of CT and X-ray cases as a whole.

**Conclusion:** Quantitative evaluation showed high accuracy for automatic diagnosis of COVID-19. Pre-trained deep learning models develop in this study could be used early screening of coronavirus, however it calls for extensive need to CT or X-rays dataset to develop a reliable application.

## 1 Introduction

The revolution of atypical and individual-to-individual transmissible pneumonia brought about by the serious intense respiratory disorder, Coronavirus (SARS-COV-2, also known as COVID-19 and 2019-nCov) has caused a global alarm [26]. In December 2019, Wuhan, Hubei province, China, turned into the focal point of an outbreak of unknown cause of pneumonia, which got attention not only in China but internationally [24]. Later Chinese Center for Disease Control and Prevention (China CDC) determined a non-SARS novel coronavirus in a patient of Wuhan at Jan 7, 2020. As of 3 April, more than 1m people have been infected in 180 countries, according to the Johns Hopkins University Center for Systems Science and Engineering.There have been over 53,000 deaths globally. Just over 3,000 of those deaths have occurred in mainland China.More than 211,000 people are recorded as having recovered from the coronavirus. [1].

Coronaviruses (CoV) are a large family of viruses that cause sickness ranging from the common cold to more severe diseases such as Middle East Respiratory Syndrome (MERS-CoV) and Severe Acute Respiratory Syndrome (SARS-CoV). Coronaviruses caused diseases in mammals and birds, a zoonotic virus that is transmitted between animals and people. Detailed investigations found that SARS-CoV was transmitted from civet cats to humans and MERS-CoV from dromedary camels to humans. When a virus spread circulating animal populations affect human population, this is termed as spillover event. It is speculated that the 2019 novel Coronaviurs was originated in bats and was transmitted in humans, possibly with pangloin as an intermediatory hosts. Coronavirus disease (COVID-19) is a new strain that was discovered in 2019 (not previously recognized in humans). Unfortunately, COVID-19 is now spreading from humans to humans.

COVID-19 diagnosis relies on clinical symptoms, positive CT images or positive pathogenic testing, and epidemiological history. Common clinical characteristics of COVID-19 include respiratory symptoms, fever, cough, shortness of breath and breathing difficulties. In more severe cases, infection can cause pneumonia, severe acute respiratory syndrome, kidney failure and even death [25, 3, 13, 9]. However, these symptoms may related to isolated cases e.g. infected CT scan of chest indicated pneumonia while pathogenic test came back positive for the virus. If a person is identified under investigation, samples of lower respiratory will be collected for pathogenic test like sputum, tracheal aspirate, and bronchoalveolar lavage etc. The laboratory technology used sequencing of nucleic acid and real time reverse-transcription polymerase chain reaction (RT-PCR) for the detection of COVID-19 [5, 4]. The detection rates of nucleic acid is low estimates (between 30% - 50%), due to the factors like availability, quality, quantity, reproducibility and stability of testing and detection kits [5, 4, 28]. Therefore, tests need to be repeated many times after confirmation in many cases.

Two well known radiological imaging modalities are used for the detection of COVID-19 i.e. X-rays and Computed Tomography (CT) scans. These two modalities are frequently used by the clinicians to diagnose pneumonia, respiratory tract infection, lung inflammation, enlarged lymph nodes, and abscesses. Since COVID-19 effect the epithelial cells of respiratory tract,thus X-rays are used to analyze the health of a patients lungs. However, most of COVID-19 cases have have similar features on CT images like ground-glass opacity in the early stage and pulmonary consolidation in the late stage. Therefore, CT scans play an important role in the diagnosis of COVID-19 as an advanced imaging modality.

Machine learning researchers and computer scientists play a vital role in the era when COVID-19 spreads all over the World. One of breakthrough of AI is Deep Learning that extracts the detailed features from the images [15]. This will help the doctors and healthcare practitioners in real time assistance, less costly, time effectively, and accurately. In this work, We have used pre-trained knowledge to improve the diagnostic performance using transfer learning techniques and compared the performance different transfer learning methods. Evaluation results using K-fold (10) showed that we have achieved state of the art performance. The contributions of researchers using machine leaning and deep learning based techniques for the detection of COVID-19 are presented in Section 2. The datasets information deployed in our study is detailed in Section 3. Then we present our proposed COVID-19 detection system based on transfer learning technique in Section 4. The evaluated results are discussed in Section 5. Finally we draw conculsion of our study in Section 6.

### 2 Literature Review

In last few months, the machine learning based techniques for identification and detection of COVID-19 have been performed. This section details the recent and current literature of COVID-19 disease detection using scan images of lungs.

Fang et al. [6] identified the viral nucleic acid for the detection of COVID-19. They compared the sensitivity of RT-PCR and non-contrast chest CT. They examined 51 patients with acute respiratory symptoms of unknown cause. The sensitivity of RT-PCR was 71% while sensitivity of non contrast chest CT was 98% for detection of COVID-19 infection. Gozes et al. [7] proposed the automated AI based system that detect the presence of COVID-19. They conducted experiment on 157 international patients of China and US. They employed 2D and 3D deep learning models to visualizes the virus effects on the lungs and achieved 98.2% sensitivity. In [26], Wang et al. conducted experiment on 1,119 CT scans of pathogen confirmed COVID-19 cases. They deployed transfer learning techniques of deep learning using Inception model and attained 89.5% accuracy.

Chen et al. [2] collected 46,096 anonymous CT images including 51 patients of confirmed COVID-19 pneumonia. They identified the COVID-19 pneumonia from CT images using UNet++ and achieved 95.24% accuracy. In [27], Xu et al. developed deep learning based system to differentiate COVID-19 pneumonia from Influenza-A viral pneumonia. They used ResNet with Location-attention classification on 618 CT samples and attained 86.7 % accuracy. Shan et al. [18] segmented COVID-19 infection regions from CT scans using deep learning model named as VB-Net. They used 249 images in traning and 300 in validation. Dice similarity coefficient of 91.6% was achieved.

Song et al. [20] presented another study investigated COVID-19 pneumonia from 51 patients of Wuhan, China. In [21], they proposed a deep neural network named Details Relation Extraction neural network (DRE-Net) to obtain the image level predictions by extracting the deep details from CT images. They used CT scans of 88 patients of COVID-19, 101 patients of bacteria pneumonia, and 86 healthy persons. They reported 94% accuracy, respectively.

### 3 Dataset

One of prerequisite in image recognition and computer vison reserch is of finding appropriate dataset. Massive amount of training dataset is one of essential requirement of deep learning, however, large medical imaging data is an obstacle in the success of deep learning. COVID-19 lung images datasets are currently limited as it is recent emerging disease. Dr. Joseph Cohen established the public GitHub repository where X-Ray and CT images of COVID-19 are collected as confirmed cases are reported. The dataset contains images of ARDS (Acute Respiratory Distress Syndrome), MERS (Middle East Respiratory Syndrome), Pneumocystis, SARS (Severe Acute Respiratory Syndrome), and Streptococcus. We have collected 200 COVID-19 images from Covid2019 chestXray dataset of GitHub repository [**?**] accessed on April 01, 2020. The metadata.csv provided with the dataset is parsed to select the positive samples of COVID-19 images. Furthermore, we collect 200 images of bacterial and viral pneumonia from Kaggle repository entitled by Chest X-Ray Images (Pneumonia) [**?**]. In this way, dataset of 200 X-Ray and CT images of COVID-19, 200 images of healthy subjects, 200 images of patients affected from bacterial pneumonia, and 200 images of patients affected from viral pneumonia respectively.

### 4 Proposed COVID-19 Detection System

Recently several deep learning methods have been proposed for the diagnosis of COVID-19. In order to improve the diagnostic performance, we have used transfer learning for diagnosis of corona virus through the transfer of already learnt knowledge. For this purpose, we used ImageNet as a source domain denoted by *D*_*s*_ where the source task *T*_*s*_ is to classify 1000 classes of natural images. The target domain denoted by *D*_*t*_ is the Covid2019 Chest X-Ray dataset where the target task *T*_*t*_ is to classify two classes i.e. Covid2019 and Healthy in binary classification and four classes i.e. Covid2019, healthy, bacterial pneumonia, and viral pneumonia in multi classification. The source domain consist of two components:

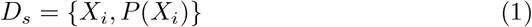

where *X*_*i*_ is the feature space of ImageNet and *P* (*X*_*i*_) is the marginal distribution such that *X*_*i*_= *{x*1, …, *x*_*n*_*}*, *x*_*i*_ *∈ X*_*i*_. The target domain also consists of two components:

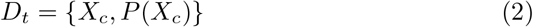

where *X*_*c*_ is the feature space of Covid2019 Chest X-Ray dataset and *P* (*X*_*i*_) is the marignal distribution such that *X*_*c*_=*{x*_1_, …, *x*_*n*_*}, x*_*j*_ *∈ X*_*c*_. On the other hand, source task *T*_*s*_ is the tuple of two elements defined as:

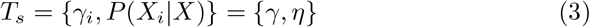

where *γ* is the label space of source data and *η* is the objective function that learned from the pair of feature vector and label space i.e.(*x*_*i*_, *y*_*i*_) *x*_*i*_ *∈ X*_*i*_, *y*_*i*_ *∈ γ*_*i*_.

The target task *T*_*t*_ is the tuple of two elements defined as:.

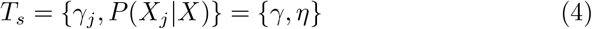

where *γ* is the label space of source data and *η* is the objective function that learned from the pair of feature vector and label space i.e.(*x*_*j*_, *y*_*i*_) *x*_*j*_ *∈ X*_*j*_, *y*_*j*_ *∈ γ*_*j*_.

The objective of transfer learning is to train a base network on large source dataset and then transfer learned features to the small target dataset. For the given source domain *D*_*s*_ and target domain *D*_*t*_ with their corresponding source task *T*_*s*_ and target task *T*_*t*_, the aim of transfer learning is to learn conditional probability distribution of target domain i.e. (P*Y*_*T*_ |*X*_*T*_) with respect to the information gain of source domain *D*_*s*_ and source task *T*_*s*_.

The proposed system employs different powerful CNN architectures (AlexNet, SqueezeNet, GoogLeNet, VGG, MobileNet, ResNet18, ResNet50, ResNet101, and DenseNet) coupled with transfer learning techniques. The system consists of three main steps: Image Acquisition, Off the shelf pre-trained models as feature extractors, and classification as illustrate from Figure 1. In the first step, images are acquired from two well known dataset: Covid2019 Chest X-Ray downloaded from GitHub repository and Chest X-Ray Images (Pneumonia) from Kaggle repository. The next step is to extract the automated features using off the shelf pre-trained architectures of CNN. In the last step, two types of classification is performed: binary classification and classification. In binary classification three scenarios are investigated: COVID-19 and Healthy classification, COVID-19 and bacterial pneumonia classification, and COVID-19 and viral pneumonia classification respectively. In multi-class problem, two scenarios are explored using three classes i.e. COVID-19, healthy, and bacterial pneumonia. Furthermore four classes are also used in the experimentation of multi-class classification i.e. COVID-19, healthy, bacterial pneumonia, and viral pneumonia. The detail of steps are described in the coming sections.

**Fig. 1:**
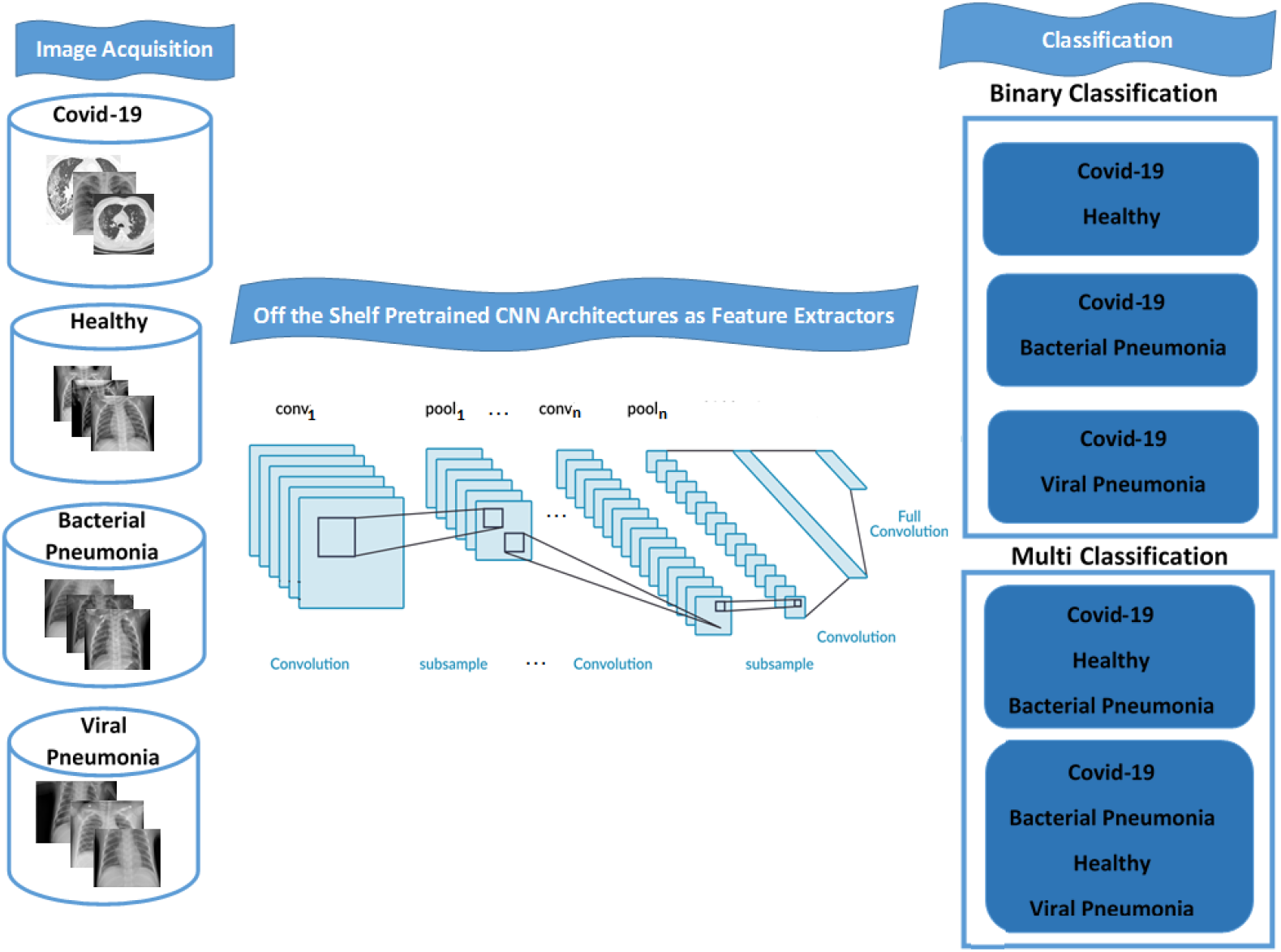
Proposed system for detection and classification of COVID-19.

### 4.1 Off the shelf pre-trained models as feature extractors

Deep learning models have got significant interest in the last few decades due to its capability of extracting automated features [14, 16]. The automated features extracted by CNN models have the capability of parameter sharing and local connectivity, thus, these features are more robust and discriminative than traditional features. The layered architecture of CNN is used to extract the features from different layers. Initial layers contain generic features (lines, edges, blobs etc.) while later layers contain specific features. The purpose of fine-tuning is to extract the features from pre-trained CNN architectures and then train a shallow network on these features. The key idea is to leverage the weighted layers of pre-trained model to extract features of ImageNet but not to update the weights of the model’s layers during training with data for the detection of COVID-19.

We used seven powerful pre-trained architectures of CNN (AlexNet, VGG, SqueezeNet, GoogLeNet, MobileNet, ResNet with its variants, and DenseNet) for extracting the robust automatic features. Utilizing these CNN models without its own classification layer enable us to extract features for our target task based on the knowledge of source task.

**AlexNet** [12] was proposed by the Supervision group, in which members were Alex Krizhevsky, Geoffrey Hinton, and Ilya Sutskever. The architecture contains 5 convolutional layers (Conv1 with 96 filters of 11*×*11, Conv2 with 256 filters of 5*×*5, Conv3 and Cov4 with 384 filters of 3*×*3, and Conv5 with 256 filters of 3*×* 3), three fully connected layers (FC6 and FC7 with 4096 neurons and FC8 with 1000 classes of ImageNet dataset).

**VGG** [19] has a uniform architecture designed by Simonyan and Zisserman. It comprises 16 convolutional layers with 3 *×*3 convolutions (but with lot of filters) and three fully connected layers like AlexNet.

One of micro-architecture of CNN was released in 2016 named as **SqueezeNet** [11]. It contains fire modules with squeezed convolutional layer(with 1 *×* 1 filters) fallow with an expanded layer (with a combination of 1 *×*1 and 3*×* 3 filters).

**GoogLeNet** [22] was proposed by Szegedy et al. in 2014. The concept of inception module was introduced in GoogLeNet in which different convolutional and pooling layers are concatenated for improving learning. GoogLeNet contains 9 inception modules, 4 max-pooling layers, 2 convolutional layers, 1 convolutional layer for dimension reduction, 1 average pooling, 2 normalization layers, 1 fully connected layer, and finally a linear layer with softmax activation in the output.

**ResNet** [8, 23] is short name of residual network in which the idea of skip connection was introduced. Skip connection means stacking the convolution layers together one after the other to mitigate the problem of vanishing gradient. Three variants of ResNet used in this study are: ResNet18, ResNet50, and ResNet101. **ResNet18** contains of 5 convolution blocks, each contain 2 residual blocks. Each residual block contains 2 convolution layers with the same number of 3 *×* 3 filters. The **ResNet50** contains 5 residual blocks each with a convolution and identity block. The convolution blocks and identity blocks has 3 convolution layers. Similarly, **ResNet101** contains 3 residual blocks with 3 convolution and identity blocks.

The idea of skip connections has been extended to connect the later blocks of densely connected layers in **DenseNet** [10] model. The dense blocks contains 1 *×* 1 convolutional filters and max-pooling layers in order to reduce the number of tunable parameters. Contrary to the skip connections in the ResNets, the output of the dense block is not added but instead concatenated. **MobileNetV2** [17] have a new CNN layer, the inverted residual and linear bottleneck layer. The new CNN layer builds on the depth-wise separable convolutions. The detail of pretrained architectures employed in this study are presented in Table 2

**Table 1:** Summarised the employed machine learning based studies for COVID-19 detection.

| References | Dataset | Study proposal | Results |
| --- | --- | --- | --- |
| Fang et al. [6] | 51 patients with acute respiratory symptoms of unknown cause | Compared the sensitivity of RT-PCR and non-contrast chest CT | Sensitivity of 71% for RT-PCR and 98% for CT |
| Gozes et al. [7] | 157 international patients of China and US | Proposed the automated AI based system that detect the presence of COVID-19 by employing 2D and 3D deep learning models | 98.2% sensitivity |
| Wang et al. [26] | 1,119 CT scans of pathogen confirmed COVID-19 cases | Deployed transfer learning techniques of deep learning using Inception model | 89.5% accuracy |
| Chen et al. [2] | 46,096 anonymous CT images | Identified the COVID-19 pneumonia from CT images using UNet++ | 95.24% accuracy |
| Xu et al [27] | 618 CT samples | Developed deep learning based system to differentiate COVID-19 pneumonia from Influenza-A viral pneumonia using ResNet with Location-attention classification | 86.7 % accuracy |
| Shan et al. [18] | 249 CT images in training and 300 in validation | Perform segmentation of COVID-19 infection regions from CT scans using deep learning model named as VB-Net | 91.6% Dice similarity coefficient |
| Ying et al. [21] | CT scans images (88 patients of COVID-19, 101 patients of bacteria pneumonia, and 86 healthy persons) | Employ Details Relation Extraction Neural network (DRE-Net) to obtain the image level predictions of corona-19 virus form healthy and bacterial pneumonia | 94% accuracy |

**Table 2:** Summary of Pre-trained CNN Architectures Employed in Proposed System

| Pre-trained Model | Input Size | Layers | Size (MB) | Parameters (M) |
| --- | --- | --- | --- | --- |
| Alexnet | $227 \times 227$ | 8 | 240 | 60 |
| SqueezeNet | $227 \times 227$ | 18 | 4.8 | 5 |
| GoogLeNet | $224 \times 224$ | 22 | 96 | 6.8 |
| VGG16 | $224 \times 224$ | 16 | 528 | 138 |
| MobileNetv2 | $224 \times 224$ | 53 | 16 | 3.5 |
| ResNet18 | $224 \times 224$ | 18 | 45 | 11.174 |
| ResNet50 | $224 \times 224$ | 50 | 98 | 23.52 |
| ResNet101 | $224 \times 224$ | 101 | 170 | 42.512 |
| DenseNet201 | $224 \times 224$ | 201 | 80 | 20.2 |

### 4.2 Classification and Detection

After extracting the automated features, we again employ pretrained CNN architectures for classification and detection of COVID-19. The pre-trained architectures used in our study for classification are: AlexNet, VGG, SqueezeNet, GoogLeNet, MobileNet, ResNet with its variants, and DenseNet. Each of these networks have three fully connected (FC) layers, where the last fully connected is used for the classification purpose. We initialize the number of neurons of last fully connected layer according to the target dataset.

We train the network on Caffee library 7 with GPU (NVIDIA CUDA) having the multiple processors of 2.80 GHz, 16GB DDR4-SDRAM, 1TB HDD, 128 GB SSD. The parameters of fine-tuning method are not set by the network itself, and it is essential to set and optimize these parameters according to the results of training the images in improving the performance. In our case, each network is trained with Adam optimizer in maximum 30 epochs. The value of batch size is set to 11 with the initial learn rate of 3*e*^*−*^4. The number of best epochs is varied according to validation criteria with validation frequency.

Two types of classification has been performed in our study: binary classification and multi-class classification. In binary classification, the dataset is split into two categories: COVID-19 and Healthy, COVID-19 and bacterial pneumonia, and COVID-19 and viral pneumonia. The images of healthy lungs are acquire from Kaggle repository, while images of lungs affected from COVID-19 are taken from GitHub repository. We will not only classify the healthy lungs from COVID-19, but also classify different causes of pneumonia either cause by some bacteria, virus, or COVID-19. Thus in multi-class classification, dataset is split into 3 categories: Healthy, COVID-19, Pneumonia-Bacterial and 4 categories: Healthy, COVID-19, Pneumonia-Bacterial, and Pneumonia-Viral. Like binary classification, images of COVID-19 are taken form GitHub repository, while images of healthy, pneumonia bacterial, and pneumonia viral are acquaint from Kaggle repository. The detail of both scenarios along with results are presented in coming section.

### 5 Result analysis and Discussions

In this section, we analyze the effectiveness of our proposed framework in light of results of experiments conducted. As discussed earlier, the experimental study is conducted using two publicly available repositories. The statistics of datasets are presented in Table 3. We deploy CNN architectures coupled with fine-tuning strategy by splitting 80% dataset in training set, 20% of training data in validation set, and 20% in testing.

**Table 3:** Dataset Split Statisitics

| Class | BinaryClass Dataset |  |  | Multiclass Dataset |  |  |
| --- | --- | --- | --- | --- | --- | --- |
|  | Train Set | Validation Set | Test Set | Train Set | Validation Set | Test Set |
| COVID-2019 | 160 | 40 | 40 | 160 | 40 | 40 |
| Healthy | 160 | 40 | 40 | 160 | 40 | 40 |
| Bacterial Pneumonia | - | - | - | 160 | 40 | 40 |
| Viral Pneumonia | - | - | - | 160 | 40 | 40 |

The effectiveness of proposed system is evaluated by computing four major outcomes of evaluation measures: true positives (tp), false positives (fp), true negatives (tn), and false negatives (fn). We use a well known evaluation measure i.e. accuracy. **Accuracy** is used to determine the classes of proposed system correctly. To evaluate the accuracy of a test set, we compute the proportion of true positive and true negative in all evaluated cases computed as:

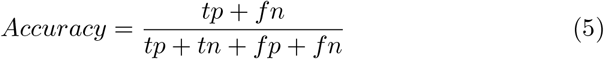

We first compute the accuracy of the proposed system for all classes either binary-class or multi-class. Then we compute mean classification accuracy using K-fold measure. Mean classification accuracy is calculated by taking average of the accuracy achieved by each of the 10 fold.

We have conduct two studies of classification in our proposed system: *Binary classification* and *Multi classification*. In binary classification, three scenarios are used: COVID-19 and healthy, COVID-19 and bacterial pneumonia, COVID-19 and viral pneumonia. Series of experiments are conducted in each scenario using nine pre-trained CNN networks i.e. AlexNet, SqueezeNet, GoogLeNet, VGG16, MobileNet, ResNet18, ResNet50, ResNet101, and DenseNet. Our initial claim is to explore the finetuning technique of transfer learning by extracting the features of pretrained CNN networks. We have achieved highest accuracy of 98.75% using VGG16, ResNet18, ResNet50, ResNet101, and DenseNet in Scenario A: COVID-19 and healthy. Similarly, we attained 98.75% using all pretrained models in Scenario B: COVID-19 and bacterial pneumonia.

The accuracy of 98.75% is achieved using SqueezeNet, GoogLeNet, MobileNet, ResNet50, and ResNet101 in Scenario C: COVID-19 and viral pneumonia. For a deeper insight on Table 4, it is observed that the performance of the automated features extracted from each pre-trained networks used in this study is comparable with one another. This supports our initial claim that fine-tuning the pre-trained CNN networks can be successfully deployed to a limited class dataset even without augmentation.

**Table 4:** Experimental Results of Binary classification and Multi classification

| Scenario | Model | Train set (%) | Validation set (%) | Test set (%) | Covid19 | Healthy (%) | Bacterial Pneumonia (%) | Viral Pneumonia (%) |
| --- | --- | --- | --- | --- | --- | --- | --- | --- |
| A. | AlexNet | 98.44 | 97.20 | 97.04 | 92.59 | 100 | — | — |
|  | SqueezeNet | 100 | 98.80 | 98.89 | 100 | 97.5 | — | — |
|  | GoogLeNet | 100 | 100 | 98.15 | 96.29 | 100 | — | — |
|  | VGG16 | 100 | 100 | 98.75 | 100 | 97.5 | — | — |
|  | MobileNet | 98.80 | 96.30 | 96.30 | 100 | 97.5 | — | — |
|  | ResNet18 | 100 | 100 | 98.75 | 100 | 97.5 | — | — |
|  | ResNet50 | 100 | 100 | 98.75 | 100 | 97.5 | — | — |
|  | ResNet101 | 100 | 100 | **98.75** | 100 | 97.5 | — | — |
|  | DenseNet | 100 | 100 | 98.75 | 100 | 97.5 | — | — |
| B. | AlexNet | 100 | 100 | 98.75 | 100 | — | 97.5 | — |
|  | SqueezeNet | 100 | 100 | 98.75 | 100 | — | 97.5 | — |
|  | GoogLeNet | 100 | 100 | 98.75 | 100 | — | 97.5 | — |
|  | VGG16 | 100 | 100 | 98.75 | 100 | — | 97.5 | — |
|  | MobileNet | 100 | 100 | 98.75 | 100 | — | 97.5 | — |
|  | ResNet18 | 100 | 100 | 98.75 | 100 | — | 97.5 | — |
|  | ResNet50 | 100 | 100 | 98.75 | 100 | — | 97.5 | — |
|  | ResNet101 | 100 | 100 | **98.75** | 100 | — | 97.5 | — |
|  | DenseNet | 100 | 100 | 98.75 | 100 | — | 97.5 | — |
| C. | AlexNet | 98.14 | 97.01 | 92.54 | 81.48 | — | — | 100 |
|  | SqueezeNet | 100 | 100 | 98.51 | 100 | — | — | 97.5 |
|  | GoogLeNet | 100 | 100 | 98.51 | 100 | — | — | 97.5 |
|  | VGG16 | 100 | 89.55 | 88.06 | 96.55 | — | — | 82.5 |
|  | MobileNet | 100 | 100 | 98.51 | 97.5 |  |  | 100 |
|  | ResNet18 | 100 | 100 | 95.52 | 92.59 |  |  | 97.5 |
|  | ResNet50 | 100 | 100 | 98.75 | 100 |  |  | 97.5 |
|  | ResNet101 | 100 | 100 | **98.75** | 100 |  |  | 97.5 |
|  | DenseNet | 100 | 100 | 97.01 | 96.29 |  |  | 97.5 |
| D. | AlexNet | 100 | 100 | 87.85 | 96.29 | 77.5 | 92.5 | — |
|  | SqueezeNet | 100 | 100 | 94.39 | 100 | 97.5 | 87.5 | — |
|  | GoogLeNet | 100 | 100 | 91.59 | 96.29 | 85 | 95 | — |
|  | VGG16 | 98.37 | 98.37 | 92.52 | 100 | 95 | 85 | — |
|  | MobileNet | 100 | 100 | **97.20** | 100 | 100 | 92.5 | — |
|  | ResNet18 | 99.77 | 99.77 | 95.33 | 100 | 92.5 | 80 | — |
|  | ResNet50 | 100 | 100 | 92.52 | 100 | 90 | 90 | — |
|  | ResNet101 | 100 | 100 | 89.72 | 100 | 92.5 | 80 | — |
|  | DenseNet | 100 | 99.77 | 93.46 | 100 | 87.5 | 95 | — |
| E. | AlexNet | 99.81 | 99.32 | 63.27 | 92.59 | 55 | 55 | 60 |
|  | SqueezeNet | 96.26 | 94.56 | 70.07 | 92.59 | 87.5 | 45 | 57.5 |
|  | GoogLeNet | 100 | 100 | 75.51 | 100 | 85 | 75 | 50 |
|  | VGG | 97.96 | 97.11 | 63.95 | 92.59 | 62.5 | 65 | 45 |
|  | MobileNet | 99.83 | 100 | **80.95** | 96.29 | 87.5 | 75 | 70 |
|  | ResNet18 | 100 | 100 | 74.15 | 100 | 70 | 80 | 55 |
|  | ResNet50 | 99.32 | 99.32 | 72.79 | 100 | 70 | 77.5 | 52.5 |
|  | ResNet101 | 98.98 | 97.96 | 70.75 | 92.59 | 77.5 | 60 | 60 |
|  | DenseNet | 96.26 | 94.56 | 70.07 | 92.59 | 80 | 70 | 45 |

The study of multi-class classification is further divided into two scenarios. In Scenario D, dataset is split into three classes: COVID-19, healthy, and bacterial pneumonia. We achieve 97.20% in this scenario using MobileNet. In Scenario E, dataset is split into four categories: COVID-19, healthy, bacterial pneumonia, and viral pneumonia. We achieve the highest accuracy of 80.95% using MobileNet architecture of CNN on test set. It can be observed from Table 4 that accuracy on multi-class problem ranging from 63.95 to 80.95. To get deeper insight, class-wise accuracy is reported in Table 4. It can be noted from Table 4 that some of bacterial and viral pneumonia leads to misclassification. The proposed system is classifying most of the viral pneumonia scans as healthy person.

For more illustration confusion matrix of each scenario with model of highest accuracy is presented in Figure 2. The scenario A accurately classify Covid19 with 100% classification rate and misclassify one instance of healthy subject with overall test accuracy of 98.75%. The scenario B and C classify Covid19 from bacterial pneumonia and viral pneumonia with 98.75% classification rate using ResNet101. In multiclassification scenarios, it is noted that viral pneumonia and bacterial pneumonia is classify as healthy subject. It is due to reason that COVID-19 is cause by SARS-COV-2 virus and pneumonia is also cause by virus or bacteria thus we need a strong overlap in the feature space between these two classes. The main reason is as the immunity system of human being has got adaptation towards pre-existing viruses. So, the human being when get infected by old viral pneumonia then the they did not have so many in the the chest x-rays or CT scans. Number of viral pneumonia is mis-classified as a bacterial pneumonia. Another reason is the pediatric x-ray images and virus attacked the child for the first time after the birth. Sometimes, the proposed system mis-classified some images of bacterial pneumonia as COVID-19 and vice versa. In case of COVID-19, when a person get pneumonia then it shows alot changes in the x-rays or CT like bacteria pneumonia or worst because human being has not yet developed the immunity against this novel corona virus. The COVID-19 images are classified accurately by number of architectures of CNN. The accuracy will be increase with increased data, more deep mesoscopic model, and long finetuning transfer learning process.

**Fig. 2:**
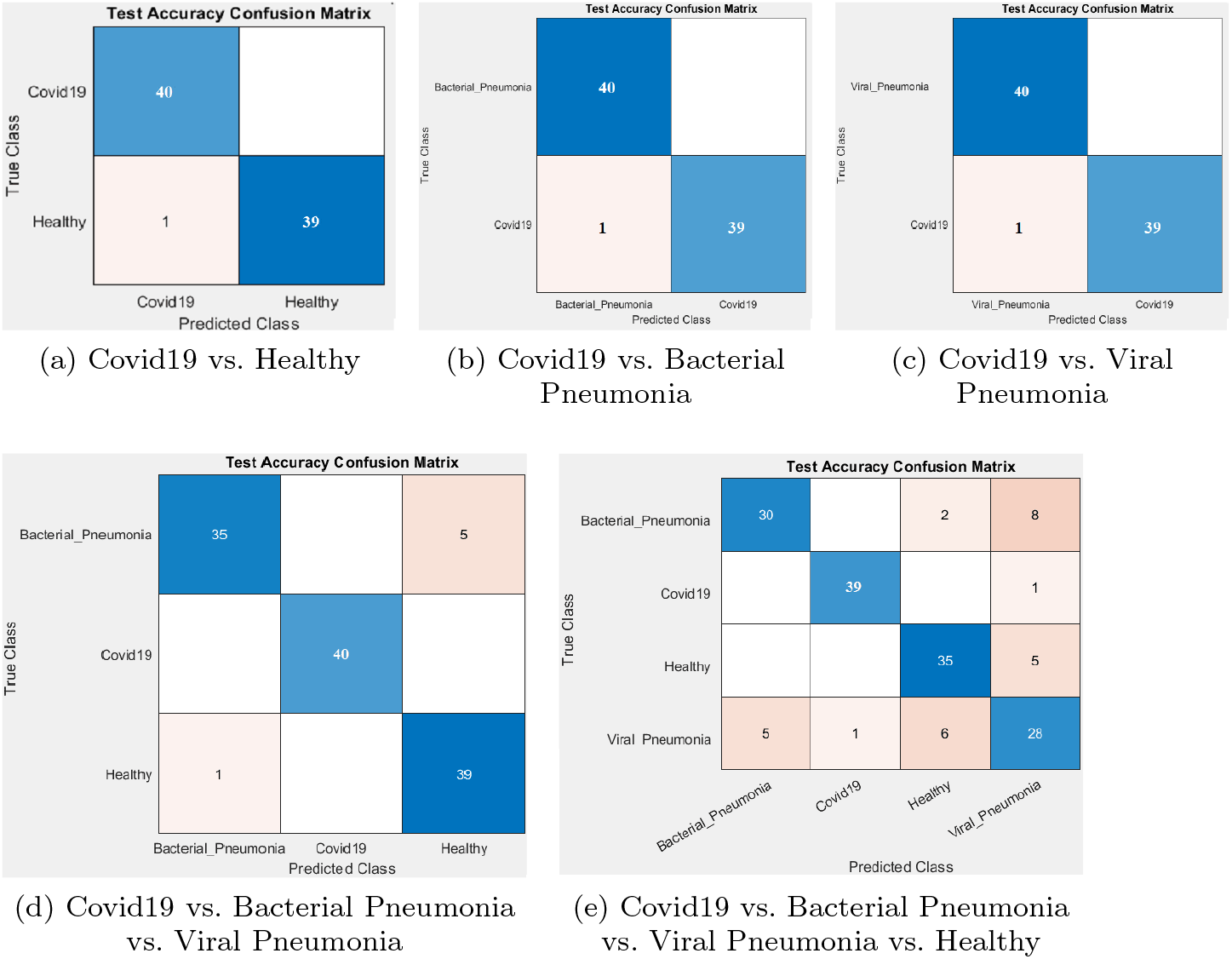
Analysis of highest identification rate of each class using different CNNs architectures: (a), (b), (c) ResNet101 (d), (e) MobileNet.

In addition with accuracy, we further evaluate our proposed system with four evaluation measures: sensitivity, specificity, precision and F-score. Sensitivity or Recall measure the ability of system to correctly classify the classes and is calculated from the proportion of true positives. It is calculated as:

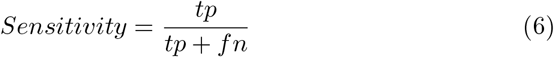

Specificity is the ability of the model to accurately classify the actual class and is computed as:

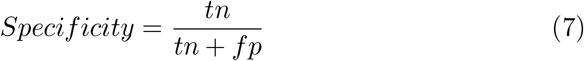

Precision is the true positive relevant measure and is calculated as:

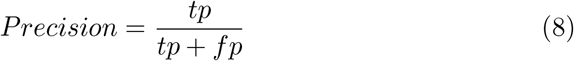

F-score or *F*_1_ score or F-measure is used to measure the accuracy of test set. It is the harmonic mean of precision and recall measured as:

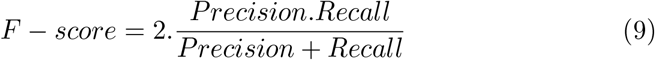

Overall accuracy, sensitivity, specificity, precision and F-score of proposed system achieved by employment of each CNN architecture using fine-tuned features with 20% test set in both scenario is presented in Table 5.

**Table 5:** Overall accuracy of COVID-19 detection system achieved by employment of each CNN architecture using fine-tuned features (20% Test set)

| Exp | Model | Accuracy | Specificity | Sensitivity | Precision | F-Score |
| --- | --- | --- | --- | --- | --- | --- |
| 1 | AlexNet | 96.30 | 0.9259 | 100 | 93.10 | 96.42 |
| 2 | SqueezeNet | 98.15 | 100 | 96.43 | 100 | 96.30 |
| 3 | GoogLeNet | 98.15 | 100 | 96.43 | 100 | 96.30 |
| 4 | VGG | 98.75 | 97.50 | 100 | 96.43 | 98.18 |
| 5 | MobileNet | 96.30 | 100 | 92.59 | 100 | 96.15 |
| 6 | ResNet18 | 98.75 | 97.50 | 100 | 96.43 | 98.18 |
| 7 | ResNet50 | 98.75 | 97.50 | 100 | 96.43 | 98.18 |
| 8 | ResNet101 | 98.75 | 97.50 | 100 | 96.43 | 98.18 |
| 9 | DenseNet | 98.75 | 97.50 | 100 | 96.43 | 98.18 |

| B. Scenario |  | COVID-19 |  | Bacterial Pneumonia |  |  |
| --- | --- | --- | --- | --- | --- | --- |
| Exp | Model | Accuracy | Specificity | Sensitivity | Precision | F-Score |
| 1 | AlexNet | 98.75 | 97.50 | 100 | 96.43 | 98.18 |
| 2 | SqueezeNet | 98.75 | 97.50 | 100 | 96.43 | 98.18 |
| 3 | GoogLeNet | 98.75 | 97.50 | 100 | 96.43 | 98.18 |
| 4 | VGG | 98.75 | 97.50 | 100 | 96.43 | 98.18 |
| 5 | MobileNet | 98.75 | 97.50 | 100 | 96.43 | 98.18 |
| 6 | ResNet18 | 98.75 | 97.50 | 100 | 96.43 | 98.18 |
| 7 | ResNet50 | 98.75 | 97.50 | 100 | 96.43 | 98.18 |
| 8 | ResNet101 | 98.75 | 97.50 | 100 | 96.43 | 98.18 |
| 9 | DenseNet | 98.75 | 97.50 | 100 | 96.43 | 98.18 |

| C. Scenario |  | Covid19 |  | Viral Pneumonia |  |  |
| --- | --- | --- | --- | --- | --- | --- |
| Exp | Model | Accuracy | Specificity | Sensitivity | Precision | F-Score |
| 1 | AlexNet | 92.54 | 100 | 81.48 | 100 | 89.80 |
| 2 | SqueezeNet | 98.51 | 97.50 | 100 | 96.43 | 98.18 |
| 3 | GoogLeNet | 98.51 | 97.50 | 100 | 96.43 | 98.18 |
| 4 | VGG | 88.06 | 82.50 | 96.30 | 78.79 | 86.67 |
| 5 | MobileNet | 98.51 | 97.50 | 100 | 96.43 | 98.18 |
| 6 | ResNet18 | 95.52 | 97.50 | 92.59 | 96.15 | 94.34 |
| 7 | ResNet50 | 98.75 | 97.50 | 100 | 96.43 | 98.18 |
| 8 | ResNet101 | 98.75 | 97.50 | 100 | 96.43 | 98.18 |
| 9 | DenseNet | 97.01 | 97.50 | 96.30 | 96.30 | 96.30 |

| D. Scenario |  | Three Classes |  |  |  |  |
| --- | --- | --- | --- | --- | --- | --- |
| Exp | Model | Accuracy | Specificity | Sensitivity | Precision | F-Score |
| 1 | AlexNet | 87.85 | 93.53 | 88.77 | 89.67 | 88.94 |
| 2 | SqueezeNet | 94.39 | 97.01 | 95.00 | 95.29 | 94.99 |
| 3 | GoogLeNet | 91.59 | 95.52 | 92.10 | 92.96 | 92.33 |
| 4 | VGG | 92.52 | 96.02 | 93.33 | 93.60 | 93.32 |
| 5 | MobileNet | 97.20 | 98.51 | 97.50 | 97.67 | 97.50 |
| 6 | ResNet18 | 95.33 | 97.51 | 95.83 | 95.85 | 98.83 |
| 7 | ResNet50 | 92.52 | 96.02 | 93.33 | 93.60 | 93.32 |
| 8 | ResNet101 | 89.72 | 94.53 | 90.83 | 91.22 | 90.80 |
| 9 | DenseNet | 93.46 | 96.52 | 94.17 | 94.32 | 94.16 |

| E. Scenario |  | Four Classes |  |  |  |  |
| --- | --- | --- | --- | --- | --- | --- |
| Exp | Model | Accuracy | Specificity | Sensitivity | Precision | F-Score |
| 1 | AlexNet | 74.15 | 91.25 | 75.95 | 76.67 | 74.33 |
| 2 | SqueezeNet | 70.07 | 89.30 | 70.65 | 71.61 | 70.10 |
| 3 | GoogLeNet | 75.51 | 91.61 | 77.50 | 76.52 | 76.81 |
| 4 | VGG | 63.95 | 87.72 | 66.27 | 66.25 | 66.11 |
| 5 | MobileNet | 80.95 | 93.48 | 82.20 | 82.52 | 82.23 |
| 6 | ResNet18 | 74.15 | 91.25 | 75.95 | 76.67 | 74.33 |
| 7 | ResNet50 | 72.79 | 90.10 | 74.90 | 73.23 | 72.43 |
| 8 | ResNet101 | 70.75 | 90.00 | 72.52 | 73.70 | 72.64 |
| 9 | DenseNet | 70.07 | 89.77 | 71.90 | 71.23 | 71.43 |

**Table 6:** Performance comparison of performance with recent works using machine learning techniques for CT and X-ray images

| Methods | Features | Model | Data samples | Performance |
| --- | --- | --- | --- | --- |
| Wang et al. [26] | Automated | Inception model | 453 | 89.5% |
| Xu et al. [27] | Automated | ResNet | 618 | 86.7 % |
| Proposed System | Automated | ResNet101 | 728 | 98.75 |

For the effectiveness of any proposed system, comparative analysis with the state of the art gives notable insight. We have compared the evaluation of our proposed system with existing technique discussed in Table 1. A meaningful comparison is possible with Wang et al. [26] and Xu et al.[27] presented in Table 6. Wang et al. [26] deployed transfer learning techniques using Inception model. They conducted experiment on 453 CT scans of pathogen confirmed COVID-19 cases and attained 89.5% accuracy. Another deep learning based system was proposed by Xu et. al. [27]. They differentiate COVID-19 pneumonia from Influenza-A viral pneumonia. They used ResNet with Location-attention classification on 618 CT samples and attained 86.7 % accuracy. Our proposed system significantly differentiate the COVID-19 from bacterial pneumonia with 98.75% and COVID-19 from viral pneumonia with 98.51% accuracy respectively. We have deployed nine pre-trained CNN networks to investigate the transfer learning techniques and conclude that fine tuning the pretrained CNN networks can be successfully deployed to a limited class dataset even without augmentation.

## 6 Conclusion

CT imaging is an efficient tool to diagnose COVID-19 and assess its. In this work, we used pre-trained knowledge to improve the diagnostic performance of COVID-19. In conclusion, our proposed study revealed the feasibility of finetuning transfer learning technique of deep learning to assist doctors to detect the COVID-19. Our system differentiate the COVID-19 from viral pneumonia and bacterial pneumonia. Our system holds great potential to improve the efficiency of diagnosis, isolation and treatment of COVID-19 patients, relieve the pressure of radiologists, and give control of epidemic. The proposed system 100% classify COVID-19 form healthy, COVID-19 from bacterial pneumonia, and COVID-19 from viral pneumonia using ResNet in binary classification. In multi-class classification, 97.20% is achieved on three classes and 80.95% on four classes using MobileNet respectively.

## Data Availability

Publicly available

